# MedZone Embedder: a framework for representation learning of Japanese secondary medical care areas from a national ICU registry, characterizing intensive care provision structure and regional vulnerability

**DOI:** 10.64898/2026.07.17.26358373

**Authors:** Kunihisa Ohno, Satoru Hashimoto

## Abstract

**Background:** In Japan, acute inpatient care is divided into approximately 335 secondary medical care areas, which serve as the basic units for planning healthcare delivery systems under the 8th National Health Care Plan. While comparisons between regions and facilities typically rely on a single risk-adjusted metric, this approach confuses differences in patient demographics with differences in the actual infrastructure of intensive care units (ICUs). This paper presents a framework — MedZone Embedder — for deriving data-driven indicators of regional structural vulnerability by mapping secondary medical care areas onto a learned similarity space, together with its working implementation. The paper sets out the concept, the method, a proof of concept, and an explicit staged validation program, rather than national empirical results.

**Framework:** Each area is represented by a feature vector consisting of aggregated values of intensive care provision indicators derived directly from the Japan Intensive Care Patient Database (JIPAD) — specifically, risk-adjusted mortality rates (standardized mortality ratios and an in-hospital composite indicator), technical efficiency, length of stay, readmission rates, case severity, and case composition — with the within-area variance of these indicators also taken into account. No hierarchical processing by facility type is performed. A contrastive autoencoder (multilayer perceptron encoder 32→16→8, symmetric decoder) is trained by self-supervised learning, using an objective function that combines reconstruction and normalized temperature cross-entropy (NT-Xent) on noise-augmented views. The resulting 8-dimensional embedding supports area searches based on cosine similarity and anomaly scoring in the embedding space (using isolation forest, Mahalanobis distance, or k-nearest-neighbor density), which is normalized to a vulnerability score ranging from 0 to 1. If deep learning libraries are unavailable, or if the number of areas is small, an alternative method using deterministic principal component analysis is employed.

**Proof of concept:** This method was implemented and deployed within an operational ICU decision support system on a managed cloud platform. The proof of concept (PoC) is structured around five secondary medical care areas within Kyoto Prefecture and runs entirely on synthetic facility-level aggregate data constructed to follow the JIPAD indicator schema; no registry data were accessed. It generated: an aggregate provision profile for each area; an area embedding space equipped with a similar-area search function; and a vulnerability ranking that identifies areas with low patient numbers and low diversity that exhibit overall poor outcomes. At this scale, the contrastive autoencoder falls back to principal component projection. The deep learning pathway has been implemented and unit testing has been completed; training and evaluation on actual registry data are pending data-use approval and the expansion of data integration.

**Planned validation:** Validation is staged. Stage 2 trains the contrastive pathway over all secondary medical care areas containing JIPAD-participating facilities, with facilities assigned to areas through authoritative ministry lists of constituent municipalities, and assesses construct validity of the vulnerability score against public structural indicators independent of the registry: ICU and HCU bed allocation, population, and geographic accessibility. Stage 3 extends coverage to all approximately 335 areas through linkage to comprehensive claims data (NDB), addressing registry participation bias and supporting policy and disaster-resilience applications.

**Conclusions:** MedZone Embedder reframes regional comparison from single-indicator ranking to structural representation: which areas are alike, and which are structural outliers. The contribution of this paper is the framework — the proposal that the intensive care provision structure of Japanese secondary medical care areas can be learned from a national outcomes registry and read through the lens of what we call institutional debt — together with a deployed implementation and a pre-specified validation program. To our knowledge, this is a candidate first application of contrastive representation learning to Japanese secondary medical care areas.

## 1. Background

Intensive care in high-income health systems is increasingly understood as a regional resource, not only as a resource of individual institutions. The relationship between case volume and outcomes, and the case for regionalizing critical care, are well established [1, 2], and the volume– outcome and specialization structure of Japanese ICUs has recently been examined at the national scale [3]. In Japan, acute inpatient care, including a large share of critical care, is planned at the level of the secondary medical care area (二次医療圏), the geographic unit within which general inpatient services are expected to be self-contained. As of September 2020, there were 335 such areas, reduced from 344 in 2016, and they remain the basic planning units for hospital-bed and physician-distribution targets under the 8th National Health Care Plan, which began in fiscal year 2024 [4].

Benchmarking across facilities or areas usually relies on a single risk-adjusted indicator such as the standardized mortality ratio (SMR). However, such indicators are difficult to interpret when provision structures differ. In the Japanese ICU registry, adult SMRs based on APACHE III-j, APACHE II, and SAPS II ranged from approximately 0.39 to 0.53, largely because of a high proportion of elective and monitoring admissions [5]. This finding prompted the development of a Japan-specific mortality prediction model [6]. A single number cannot show whether an area is served by one dominant facility type or by a mixture of types, nor how that configuration relates to outcomes. This information is central to capacity planning and disaster resilience.

The Japan Intensive Care Patient Database (JIPAD) is a national ICU registry operated by the Japanese Society of Intensive Care Medicine. It provides high-quality, severity-scored admission data and is the intended empirical basis of this framework [5]. Contrastive representation learning of geographic regions is already established in urban computing, where regions are embedded from mobility data, points of interest, and imagery for downstream socioeconomic tasks [7, 8]. One such model embeds regions and predicts social vulnerability [10], and region embeddings have also been used to delineate health service areas and to score provider shortage [9]. Two gaps remain. First, these methods characterize regions mainly by access, mobility, and shortage, rather than by the clinical provision structure recorded in a national outcomes registry. Second, to our knowledge, they have not been applied to Japanese secondary medical care areas. The present work addresses both gaps. Facility-level typology of ICUs is a distinct, related question; it is being pursued separately in an effort led by the JIPAD Working Group and is not claimed here.

### 1.1 Conceptual motivation: institutional debt

Conventional ICU benchmarking assumes that outcomes are determined primarily by patient-level severity and facility-level performance, and that fair comparison is achieved once case mix is adjusted away. We propose a complementary premise: regional provision structures accumulate constraints over time — in volume, facility composition, internal heterogeneity, and the configuration of beds and expertise — and these constraints bound the outcomes a region can achieve regardless of how well any single admission is risk-adjusted. By analogy with technical debt in software engineering, we refer to this accumulated structural constraint as institutional debt. Institutional debt is not observable as a single indicator; it manifests as a configuration. Measuring it therefore requires a representation in which whole provision configurations can be compared — which is precisely what an embedding provides (Figure 1).

**Figure 1.**
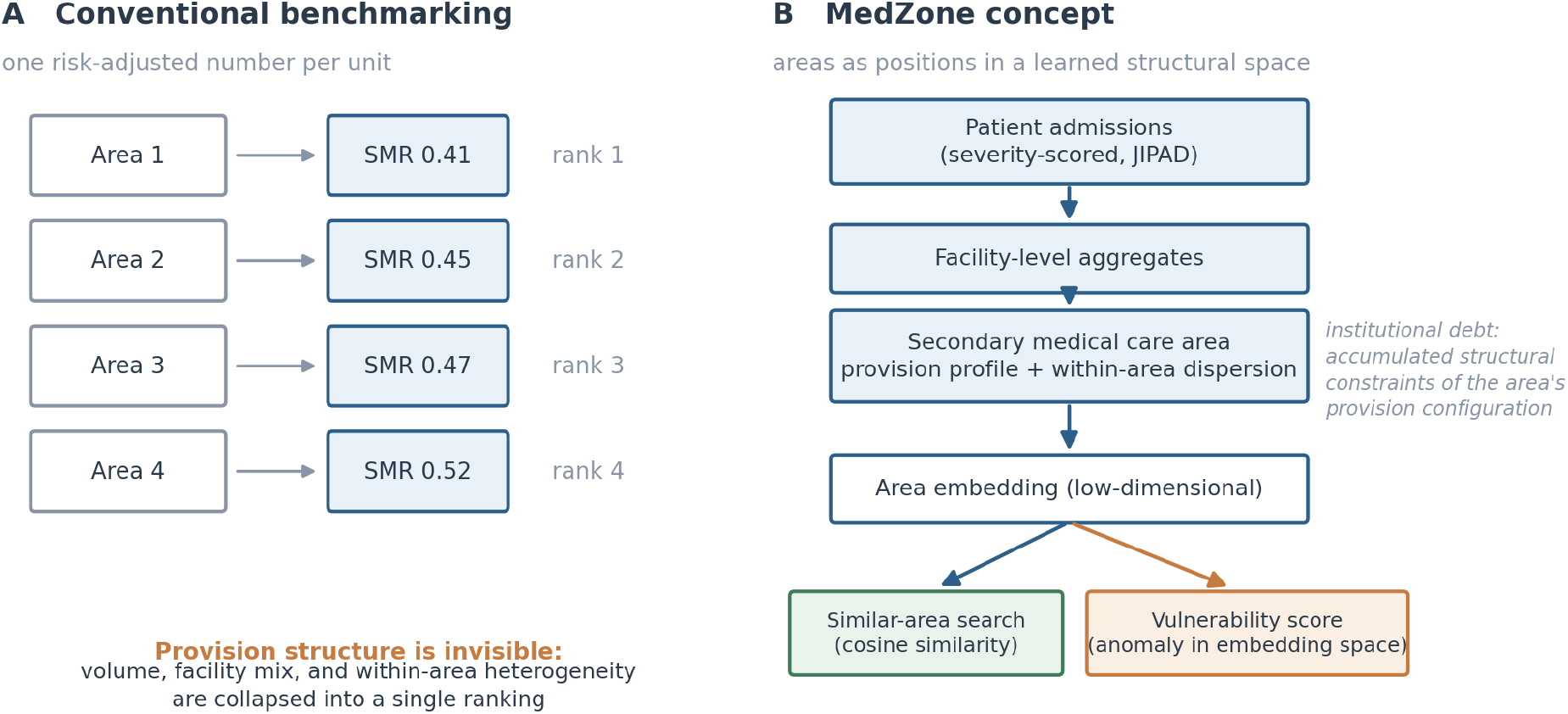
From single-indicator benchmarking to structural representation. (A) Conventional benchmarking collapses each area into one risk-adjusted number, leaving provision structure invisible. (B) The MedZone concept: severity-scored admissions are aggregated to facilities and then to secondary medical care areas, whose provision profiles (including within-area dispersion) are embedded in a learned space that supports similar-area search and anomaly-based vulnerability scoring. Institutional debt — the accumulated structural constraint of an area’s provision configuration — is what the embedding is designed to expose.

### 1.2 A layered research program

MedZone Embedder is one layer of a deliberately layered research program (Figure 2). JIPAD supplies the clinical layer: severity-scored admissions and outcomes from a national ICU registry [5]. MeshScope supplies the geographic layer: authoritative secondary-medical-care-area definitions and the bed-allocation structure of intensive care, with a route to spatial visualization of the embedding. MedZone Embedder — the subject of this paper — is the representation layer that turns area-level provision profiles into a comparable structural space. Navigator is the decision support layer through which clinicians, registries, and planners interact with the result. Separating the layers keeps responsibilities clear: data quality is owned by the registry, geography by public administrative definitions, representation by the method described here, and interpretation by the people who use it.

**Figure 2.**
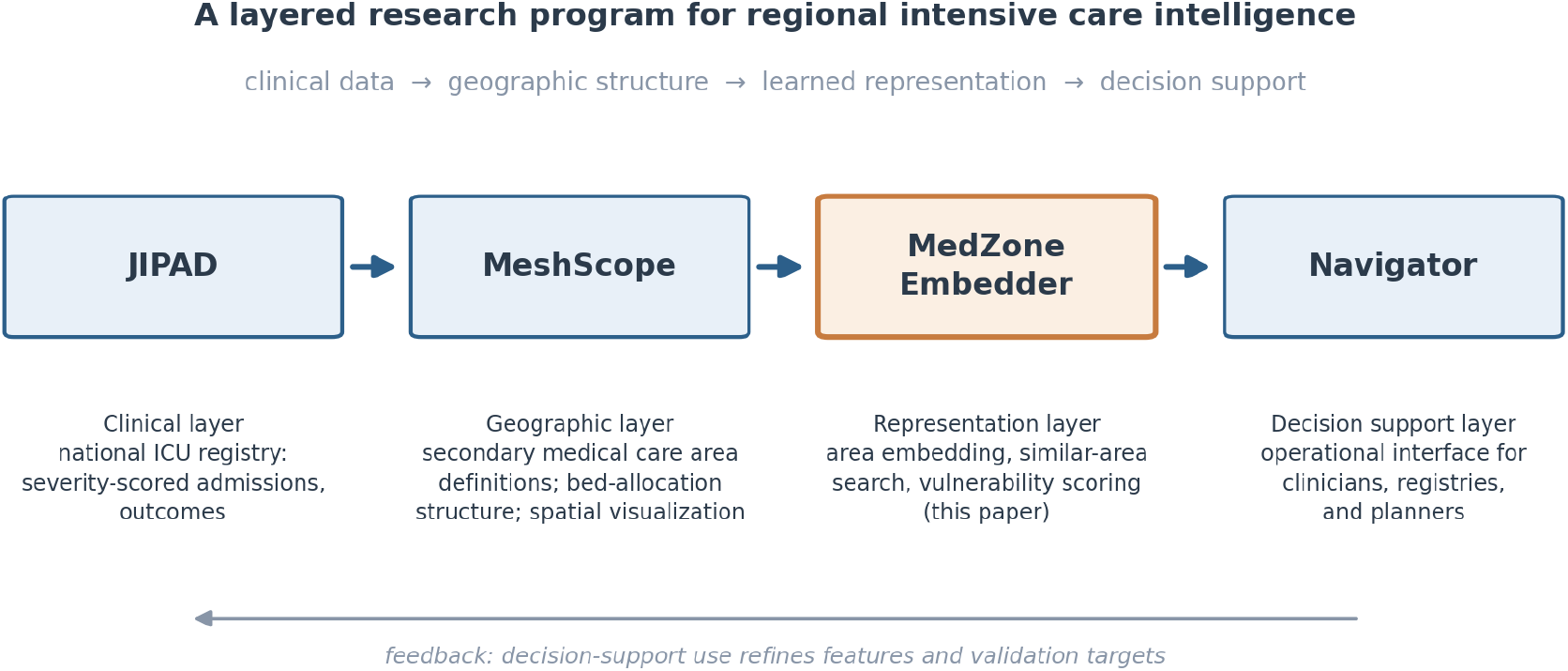
The layered research program. Clinical data (JIPAD), geographic structure (MeshScope), learned representation (MedZone Embedder; this paper), and decision support (Navigator). The feedback loop indicates that decision-support use refines features and validation targets.

### 1.3 This paper

This paper describes MedZone Embedder as a framework: a contrastive autoencoder that embeds secondary medical care areas into a low-dimensional space, supports similar-area search, and produces anomaly-based vulnerability scores, deployed within an operational decision support system. The intended contribution is the concept and the staged research program built around it, demonstrated at proof-of-concept scale; national empirical results are explicitly designated as the next stage (Section 5).

## 2. The MedZone Embedder framework

### 2.1 Setting and data

The intended empirical basis is the JIPAD registry. The currently deployed proof of concept runs the pipeline over five secondary medical care areas in Kyoto Prefecture (Kyoto·Otokuni, Yamashiro-North, Yamashiro-South, Chutan, and Tango) using synthetic facility-level aggregate data constructed to follow the JIPAD indicator schema; no actual registry data were accessed or analyzed for this demonstration. In the demonstration build, synthetic facilities are assigned to areas through a deterministic, reproducible mapping. The production target is the full set of approximately 335 national areas, analyzed with actual JIPAD data under the registry’s data-use terms and expanded data linkage (for example, to the National Database, NDB). All values shown in this report are synthetic; no individual patient records exist in the demonstration.

### 2.2 Feature construction

Each secondary medical care area is described directly by a vector of aggregate intensive care provision indicators defined over the JIPAD admissions attributed to that area (taking synthetic values in the present demonstration), without first clustering facilities into types. The indicators are: risk-adjusted mortality (the standardized mortality ratio, SMR, and a within-admission composite, STREAM-SMR); technical efficiency from data envelopment analysis; mean length of stay; readmission rate; mean APACHE III-j score and mean age as case-mix proxies; the number of contributing facilities; and the within-area dispersion (for example, the variance) of the quality and efficiency indicators. The dispersion terms allow the embedding to express provision structure — that is, whether an area’s facilities are homogeneous or heterogeneous — without producing named facility types. Facility-level typology is deliberately out of scope here and is being pursued as a separate effort led by the JIPAD Working Group; this manuscript treats the area as the unit of analysis throughout. For the national expansion, the system’s explanatory layer also anticipates NDB-era area features such as ICU supply capacity, population, and geographic accessibility.

### 2.3 Embedding model

Area vectors are standardized and embedded by a contrastive autoencoder (Figure 3). The encoder is a multilayer perceptron with hidden widths of 32 and 16 and an 8-dimensional output, using ReLU activations and dropout (0.2); the decoder is symmetric. Training is self-supervised. For each area, two views are formed by adding Gaussian noise (standard deviation 0.1), and the objective combines a reconstruction mean squared error over both views with a normalized temperature cross-entropy (NT-Xent) contrastive term,

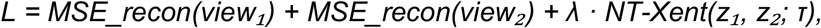

with a contrastive weight of λ = 0.5 and a temperature of τ = 0.5. Positive pairs are the two views of the same area; all other views in the batch serve as negatives. Optimization uses Adam (learning rate 1×10^−3^, weight decay 1×10^−4^) for up to 200 epochs with a batch size of 64 and early stopping (patience 20). All randomness is seeded (seed 42), and training runs on CPU. When PyTorch is unavailable, or when the number of areas is below ten, the method falls back deterministically to principal component analysis padded to the embedding dimension. This fallback is reported transparently in the output.

**Figure 3.**
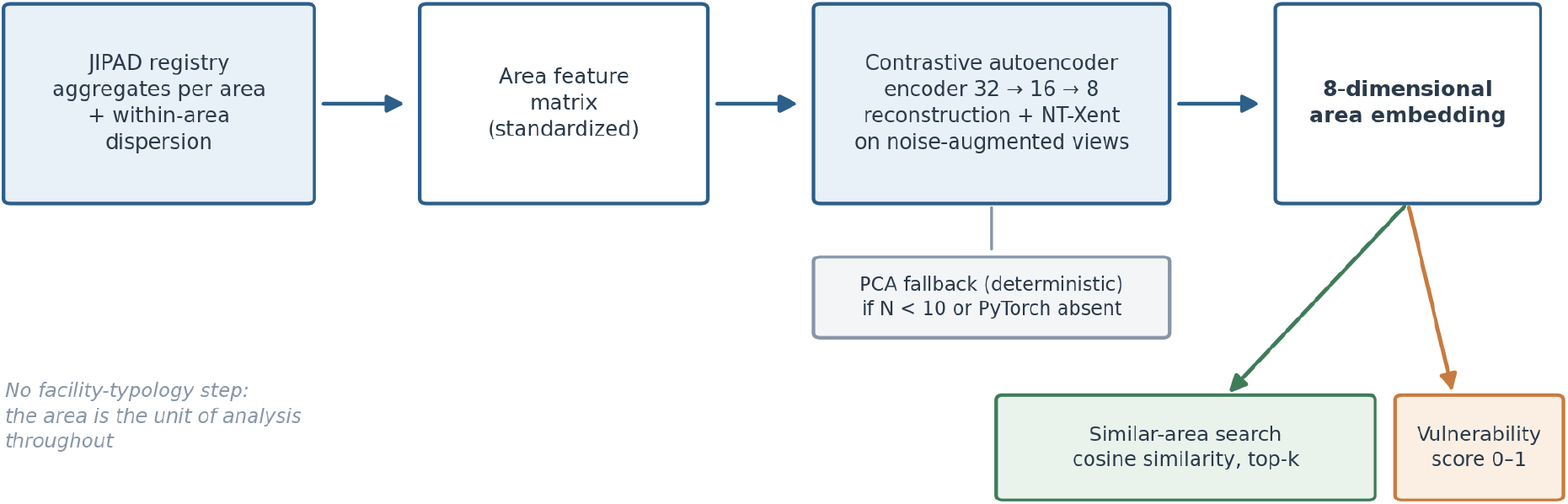
MedZone Embedder pipeline. Each secondary medical care area is described directly by aggregate intensive care provision indicators from the JIPAD registry (with within-area dispersion). These form an area feature matrix, which a contrastive autoencoder embeds into an 8-dimensional space that supports similar-area search and anomaly-based vulnerability scoring. No facility-typology step is involved; facility-level typology is a separate effort led by the JIPAD Working Group. A principal component fallback is used at small scale.

### 2.4 Configuration

**Table 1.** Embedder configuration as implemented in the core engine.

| Component | Setting | Notes |
| --- | --- | --- |
| Embedding dimension | 8 | $z \in \mathbb{R}^8$ per area |
| Encoder hidden widths | $32 \rightarrow 16 \rightarrow 8$ | ReLU; dropout 0.2; symmetric decoder |
| Augmentation | Gaussian, $\sigma = 0.1$ | two views per area |
| Contrastive term | NT-Xent, $\tau = 0.5$, $\lambda = 0.5$ | SimCLR-style objective [11] |
| Optimizer | Adam, lr $1e-3$, wd $1e-4$ | $\leq 200$ epochs, batch 64 |
| Early stopping | patience 20 | on training loss |
| Reproducibility | seed 42, CPU | deterministic |
| Fallback | PCA | if PyTorch absent or $N < 10$ |

### 2.5 Similar-area search and vulnerability scoring

Similar areas are retrieved by cosine similarity in the embedding space (top-k). Regional vulnerability is computed by anomaly detection on the embeddings, using one of three interchangeable methods: isolation forest (200 trees, contamination 0.1), Mahalanobis distance to the embedding centroid, or mean k-nearest-neighbor distance. Raw scores are min–max normalized to a vulnerability score ranging from 0 to 1 and then ranked [12]. In addition, the user interface shows a deliberately transparent heuristic baseline, vulnerability ≈ mean STREAM-SMR × (1 / facility count) × (1 / provision heterogeneity), so that areas with low volume, a narrow provision structure, and poorer outcomes receive higher scores. The heuristic score and the embedding-based score are presented side by side, rather than as a single authoritative figure.

### 2.6 Implementation and deployment

MedZone Embedder is implemented in Python and exposed through a Streamlit interface. It is deployed on Google Cloud Platform with identity-aware access control, a load balancer, and BigQuery storage. Narrative summaries are generated through Vertex AI within the platform boundary, and page access is audit-logged. The module forms part of an operational ICU decision support system. Embedding, similarity search, and vulnerability scoring are organized as a single reproducible pipeline with fixed seeds.

## 3. Proof-of-concept demonstration

The deployed proof of concept over the five Kyoto areas — run entirely on synthetic aggregate data that follow the JIPAD indicator schema — exercises the full pipeline and produces three classes of output. First, each area receives an aggregate provision profile (risk-adjusted mortality, efficiency, length of stay, readmission, case mix, facility count, and within-area dispersion), summarized in a per-area table. Second, areas are placed in an embedding space in which cosine similarity search returns the areas most structurally similar to a chosen reference area. Third, a vulnerability ranking highlights low-volume areas with poorer and more dispersed aggregate outcomes; the rank of the selected area is reported explicitly.

We emphasize the scale caveat. With five areas, the data fall below the ten-area threshold at which the contrastive autoencoder is engaged, so the deployed visualization uses the principal component fallback. The deep learning pathway is implemented and unit-tested, but it cannot be trained meaningfully at this scale, and we make no claims about outcome validation here. The proof of concept demonstrates feasibility, interpretability, and integration, not predictive performance; because the inputs are synthetic, all outputs shown are illustrative rather than empirical.

## 4. Discussion

MedZone Embedder reframes regional comparison: instead of asking “which area has a better number,” it asks “which areas are structurally alike, and which are structural outliers.” Three aspects should be distinguished. The contrastive autoencoder itself is a standard self-supervised architecture [11, 13], and contrastive embedding of geographic regions is established in urban computing [7, 8, 10]; its application to health service area delineation and provider-shortage scoring has also been demonstrated [9]. We therefore do not claim a new learning algorithm. What is new is the object and the framing: (i) Japanese secondary medical care areas are embedded by their clinical provision structure, drawn from a national outcomes registry rather than from access or mobility data; (ii) regional vulnerability is derived as an anomaly score in that learned space; and (iii) both are integrated into an operational ICU decision support system. Compared with region-embedding work built on mobility and access-based signals [7, 8, 9, 10], the registry-derived, outcome-aware characterization is the distinguishing feature. We position this work as a candidate first application in Japan, not as a world first.

The method operationalizes the layered program introduced in Section 1.2 (Figure 2): JIPAD supplies the clinical data; MeshScope supplies geographic structure and a route to spatial visualization of the embedding; and Navigator supplies the decision support interface. The institutional-debt perspective introduced in Section 1.1 motivates the design: an area’s provision configuration constrains the outcomes that can be achieved, independently of the risk adjustment applied to any single admission, and embedding makes this latent, relational structure comparable across areas. The reading of regional systems here is deliberately relational rather than essentialist: an area is characterized by its position among other areas, not by an intrinsic score.

Potential uses are hypothesis-generating. Similar-area search can identify reference areas for transferring best practice. Vulnerability ranking can suggest where backup capacity should be prioritized for disaster resilience, or where capacity planning deserves closer scrutiny. None of these uses should be treated as prescriptive on the present evidence.

## 5. Planned validation and staged research program

Because this is a framework paper, we state the validation program explicitly rather than leaving it implicit. The program is staged so that each stage has its own data requirements, its own primary question, and its own falsifiable output (Table 3).

**Table 2.** Components and honest novelty positioning, mirroring the system’s internal feature registry.

| System feature | Status | Positioning |
| --- | --- | --- |
| MedZone Embedding Engine | Implemented | Candidate first application of contrastive representation learning to Japanese secondary medical care areas |
| Regional vulnerability scoring | Implemented | Anomaly detection in the embedding space; three interchangeable methods plus a transparent heuristic |
| Facility-level typology | Out of scope | A distinct question pursued in a parallel effort led by the JIPAD Working Group; not claimed here |

**Table 3.** The staged research program. Each stage has its own data requirement, primary question, and falsifiable output.

| Stage | Scope and data | Primary question | Output |
| --- | --- | --- | --- |
| Stage 1 (this paper) | Concept; five Kyoto areas; synthetic aggregates in the JIPAD indicator schema | Is the framework coherent, implementable, and interpretable? | Framework; deployed pipeline; proof-of-concept demonstration |
| Stage 2 | All JIPAD-covered areas; authoritative facility-to-area mapping; public bed-allocation and population | Does the contrastive embedding behave at scale, and does the vulnerability score track independent | Trained national embedding; construct-validity analysis; contrastive vs. PCA |

|  | data | structural indicators? | comparison |
| --- | --- | --- | --- |
| Stage 3 | All approximately 335 areas via NDB linkage | Does full coverage change the structural picture, and what follows for planning and disaster resilience? | Participation-bias-corrected national atlas; policy applications |

Stage 2 — national training and construct validity. The analysis population is defined as all secondary medical care areas containing at least one JIPAD-participating facility. This population is expected to exceed comfortably the ten-area threshold at which the contrastive pathway is engaged, allowing the deep learning route to be trained and compared directly against the principal component fallback on identical features. Facilities will be assigned to areas through the authoritative ministry lists of constituent municipalities, replacing the synthetic data and illustrative mapping of the present demonstration. Construct validity of the vulnerability score will then be assessed against external structural indicators that are public and independent of JIPAD: ICU and HCU bed allocation by area (as compiled in the MeshScope layer from ministry data), area population, and geographic accessibility. The pre-specified expectation is that high-vulnerability areas correspond to areas with low bed supply relative to population; failure of this correspondence would count against the score as currently constructed.

Stage 3 — full national coverage and policy application. Linkage to the National Database of health insurance claims (NDB) extends coverage to all approximately 335 areas, addresses the participation bias inherent in a voluntary registry, and adds supply-side features beyond the registry’s reach. At this stage the framework can support the policy-facing questions that motivate it: where backup capacity should be prioritized for disaster resilience, and which provision configurations are associated with resilient regional outcomes.

## 6. Limitations

- The proof of concept covers only five areas and uses synthetic aggregate data that emulate the JIPAD indicator schema; no actual registry data have yet been analyzed. At this scale, the contrastive model is not engaged, and the principal component fallback is used. National-scale results are not yet available.
- Across the full set of approximately 335 areas, the low ratio of areas to features creates a risk of overfitting. The embedding dimension and hyperparameters were set by design and have not been tuned, and the Gaussian noise augmentation is generic rather than domain-informed.
- The feature definitions are currently based on JIPAD only. Because JIPAD participation is voluntary and covers a subset of Japanese ICUs, the area representations may be biased by which facilities participate. Planned NDB-era features (supply capacity, population, and accessibility) have not yet been incorporated.
- The vulnerability score has not been validated against external outcomes and should not, on its own, drive funding or closure decisions. It is presented alongside a transparent heuristic precisely to discourage over-interpretation.
- The facility-to-area mapping in the demonstration is deterministic and illustrative; real mappings require authoritative area definitions (planned for Stage 2 through ministry lists of constituent municipalities; Section 5).

## 7. Conclusions

This paper has presented MedZone Embedder as a framework: the proposal that the intensive care provision structure of Japanese secondary medical care areas can be learned from a national outcomes registry, read through the lens of institutional debt, and acted on through similar-area search and vulnerability scoring within an operational decision support system. A deployed proof of concept on synthetic data demonstrates feasibility, interpretability, and integration. The framework offers a tractable, structure-aware complement to single-indicator benchmarking, and its value will be decided by the staged program set out in Section 5: national training over JIPAD-covered areas with construct validation against independent structural indicators, followed by full national coverage through NDB linkage. We set out the framework now so that the concept, the method, and the validation criteria are public before the empirical results arrive.

## Data Availability

The demonstration in this report uses only synthetic facility-level aggregate data; no actual patient registries or human data were accessed or analyzed for this initial stage. The synthetic demonstration dataset is available from the corresponding author on reasonable request. Actual JIPAD data are available to researchers by application to the JIPAD Working Group under the JIPAD data-use terms.

## Declarations

### Ethics approval and consent to participate

The demonstration in this report uses only synthetic facility-level aggregate data; no JIPAD registry data, and no individual patient data, were accessed or analyzed. JIPAD operates under the governance of the JIPAD Working Group of the Japanese Society of Intensive Care Medicine; the planned use of registry data in Stages 2–3 will be subject to the JIPAD data-use terms and the relevant institutional review processes.

### Consent for publication

Not applicable.

### Availability of data and materials

JIPAD data are available to researchers by application to the JIPAD Working Group under the JIPAD data-use terms. The MedZone Embedder method is described in full in this manuscript; the synthetic demonstration dataset is available from the corresponding author on reasonable request. The surrounding decision support system codebase is proprietary to Jinen Co., Ltd.

### Authors’ contributions

KO conceived the method, implemented the system and the analyses, and drafted the manuscript. SH supervised the work, advised on JIPAD data governance and clinical interpretation, and critically revised the manuscript. Both authors read and approved the final manuscript.

## Acknowledgements

The authors thank the JIPAD Working Group and the participating facilities, as well as collaborating clinicians and methodologists, for their support of the registry on which this work is based.

